# What Explains the Socioeconomic Status-Health Gradient? Evidence from Workplace COVID-19 Infections

**DOI:** 10.1101/2021.03.23.21254170

**Authors:** Raphael Godefroy, Joshua Lewis

**Author notes:** We thank Marcella Alsan, Gustavo Bobonis, Abel Brodeur, Fabian Lange, Mark Stabile, and seminar partic-ipants at Université de Montréal for valuable comments and suggestions. Financial support from the Social Sciences and Research Council of Canada is gratefully acknowledged. Data can be obtained through an application process to the CIUSSS and the CISSS for the Greater Montreal Area.

## Abstract

This paper studies the contribution of the workplace to the SES-health gradient. Our analysis is based on a unique dataset that tracks various health outcomes and workplace risks among healthcare workers during the first four months of the coronavirus 2019 (COVID-19) pandemic. The setting provides an exceptional opportunity to test for work-related disparities in health, while controlling for confounding determinants of the SES-health gradient. We find that low-SES nurses were systematically more likely to contract COVID-19 as a result of workplace exposure. These differentials existed in all healthcare institutions, but were particularly large in non-hospital settings. In contrast, we find no relationship between SES and non work-related infection rates. The differences in workplace infection rates are substantially larger than those implied by standard ‘task-based’ indices of transmission risk, and cannot be attributable to easily identifiable metrics of workplace risk. Together, our results show how subtle differences in work conditions or job duties can substantially contribute to the SES-health gradient.

## 1 Introduction

The positive relationship between socioeconomic status (SES) and health is one of the most well-established findings in social science. Yet the sources of this gradient remain poorly understood. The empirical literature has focused mainly on disparities in the determinants of health, including access to medical care, risky behavior, and environmental factors both in the workplace and at home. However, because these various mediators often move in tandem, it has proven difficult to disentangle their respective contributions.

We assess the role of the workplace for the SES-health gradient among a specific population: frontline healthcare workers during the Coronavirus 2019 (COVID-19) pandemic. Our analysis draws on a dataset with information on workplace risks from COVID-19 exposure and worker health outcomes. This setting provides a unique opportunity to test for work-related disparities in health, *holding constant* all other potential determinants of the SES-health gradient.

Our analysis is based on an administrative dataset of healthcare workers in Montreal, Quebec, during the first four months of the pandemic. Montreal was a major COVID-19 hotspot during the first wave. The city had the highest rates COVID-19 cases in Canada, and had similar case rates to several major northeastern U.S. cities.^1^ The data cover healthcare workers in three different healthcare institutions: hospitals, long-term elder care facilities (CHSLDs), and local community health centers (CLSCs). There is detailed information on a range of health-related outcomes, including whether individuals had a COVID-19 contact at work, self-assessed symptoms, and diagnostic test results. There is also detailed information on occupation, age, location of work, and location of residence.

We focus on three narrowly defined occupation categories: Licensed Nurse Practitioners (LPNs), Registered Nurses (RNs), and Nurse Clinicians (NCs). These occupations each have similar daily workplace tasks – they are all nurses – but have distinct educational requirements and widely varying pay scales.^2^ For the sake of clarity, we refer to these three occupations as Low-SES, Mid-SES and High-SES nurses.

We estimate the relative probability of infection across different SES nurses who were or were not exposed to COVID-19 in the workplace. These comparisons are based on workers with similar characteristics (age, gender, and residence ZIP code) who worked in the same institution (hospital, nursing home facility, or local community health departments). The resulting estimates capture how heterogeneity in the SES-health risk gradient varies with underlying workplace conditions, holding constant all non-work drivers of COVID-19 infection. Thus, we are able to isolate the impact of the workplace on the relative risk of COVID-19 infection across different SES workers.

We find that Low-SES nurses were significantly more likely to test positive for COVID-19. This gradient is driven entirely by workplace exposure, and we find no significant difference in infection rates across nurses who were not exposed to COVID-19 in the workplace. The patterns are stable across a number of different specifications including models that control for place of work fixed effects, suggesting that the observed SES gradient cannot be attributed to differences in underlying infection risk or safety protocols across healthcare facilities.

What explains the SES gradient in workplace-related COVID-19 infection rates? One possibility is that Low-SES nurses were more likely to be tested following a workplace contact.^3^ As a result, the observed SES gradient in COVID-19 diagnoses may simply reflect a lower rate of undiagnosed infection among Low-SES nurses.^4^ We assess overall (positive plus negative) testing rates across groups. We find that Low-SES nurses who were exposed to COVID-19 at work were not significantly more likely to be tested. Moreover, the estimated SES gradient is similar when we restrict the sample to nurses who received a PCR test. Finally, we find that Low-SES nurses were significantly more likely to experience COVID-19 symptoms following a workplace contact, *regardless* of whether they were tested. Together, these findings show that differential testing rates across groups cannot account for the observed SES-infection gradient.

A second explanation is that distinct job responsibilities contributed to the observed SES-gradient. Although workers in the three occupations share similar daily routines and job duties, it is possible that the particular tasks required of Low-SES nurses placed them at heightened risk of infection. Indeed, we find that Low-SES nurses were more likely to have contact with a COVID-19 patient. Conditional on workplace contact, however, we find no significant differences in the duration of exposure or the probability of a high-risk contact. These findings suggest that basic measures of workplace risk may not capture the subtle ways in which specific job responsibilities and work conditions contributed to the differential rates of infection across nurses.

A final possibility is that the results reflect behavioral differences in the workplace. SES-based differences in adherence to safety protocols could arise from different levels of knowledge about the disease and the sources of transmission (Lange, 2011) or heterogeneous preferences to-wards risk (Cutler and Lleras-Muney, 2010). Although we cannot entirely rule out this channel, several pieces of evidence contradict a behavioral mechanism. First, unlike the general public, who may have misperceived the risks from COVID-19 early in the pandemic (Alsan et al., 2020; Simonov et al., 2020), frontline healthcare providers were more cognizant of the risks of the disease and the potential sources of infection. Second, we find no systematic differences in infections rates among nurses who were not exposed to COVID-19 at work, suggesting similar levels of risk-taking outside the workplace. Finally, we find systematic differences in the magnitude of the SES-infection gradient across different healthcare institutions, with effect sizes that are three times larger in non-hospital settings. These heterogeneous effects are inconsistent with a simple behavioral mechanism, which should affect risk-taking similarly across institutions. Instead, the patterns point to heterogeneity in the underlying level of risk across healthcare institutions. Indeed, a report by Quebec’s ombudsman found that the province focused heavily preparing hospitals for the pandemic at the expense of long-term care facilities. The report documented acute shortages of personal protective equipment (PPE) and lack of infection control measures in these institutions (Rinfret, December 10, 2020). Our results suggest that the costs of these inadequate safety measures were disproportionately borne by Low-SES nurses.

Our results complement a number of recent studies that use occupation-based measures of viral transmission risk during the COVID-19 pandemic. These measures have been used to measure the size and characteristics of at-risk populations (Baylis et al., 2020*b*; Baker, Peckman and Seixas, 2020), to evaluate differential economic effects across workers (Beland et al., 2020; Beland, Brodeur and Wright, 2020), and to guide the policy response (Aum, Lee and Shin, 2020; Chopra, Devereux and Lahiri, 2020). To measure viral transmission risk, researchers typically combine O*NET data that provide detailed information on a range of specific job requirements (such as physical proximity, frequency of face-to-face interactions, etc.) with assessments from health experts on the role of these factors for viral transmission risk (see Baylis et al., 2020*a*; Beauregard, Connolly and Haeck, 2020). Although the ordering of the O*NET ‘task-based’ measure of infection risk aligns with our estimates of observed workplace infection risk, it drastically understates the disparities in infection risk across occupations and does not account for the large differences in infection risk across healthcare institutions. Taken together, our results show how large disparities in workplace infection risk can arise, even across workers in similar occupations with similar tasks. The findings provide a cautionary note for subjective task-based measures of infection risk, and highlight the critical need for data on incidence of workplace COVID-19 infections across a variety of occupations and industries.

More broadly, our research provides new insights into the drivers of the socioeconomic health gradient. These health disparities have been widely documented in general, and have only intensified during the current pandemic (Golestaneh et al., 2020; Chowkwanyun and Reed, 2020; Baena-Diez et al., 2020). Researchers have documented a number of mediating factors in this relationship including differences in risky behavior (Grimard and Parent, 2007; de Walque, 2007; Cutler and Lleras-Muney, 2010; Cutler et al., 2011; Kenkel, 1999; Alsan et al., 2020), access to healthcare (Currie and Gruber, 1996; Goodman-Bacon, 1996; Clay et al., 2020), and environmental conditions both at home and in the workplace (Evans and Kantrowitz, 1996; Currie and Stabile, 2003; Wu et al., 2020).^5^ Nevertheless, since these plausible mechanisms operate simultaneously, it is often challenging to isolate their respective influences. Our findings suggest that on-the-job conditions may represent an important driver of the health gradient.

## 2 Data

We obtained an administrative dataset from the Human Resources department for all nurses across a subset of healthcare facilities in Montreal, Quebec from March 15 to July 1, 2020. These data provide information on workers at three types of healthcare facilities: hospitals, CHSLDs, and CLSCs. CHSLDs include both public and private residential and long-term care facilities. These centers provide temporary or permanent lodging primarily to elderly residents who are unable to live autonomously. Services include living assistance, support, and monitoring, as well as nursing, pharmaceutical, medical care, and rehabilitation. CLSCs are local community health centers run by the provincial government that service residents in a defined catchment area. Typical services include non-urgent healthcare and social services, preventative medical services, rehabilitation, and public health activities.

We focus on three distinct categories of healthcare workers: Licensed Nurses Practitioner (Low-SES), Registered Nurses (Mid-SES), and Nurse Clinicians (High-SES).^6^ These three occupations are regulated by the provincial government, each with specific educational requirements and salary profile.^7^ The educational requirements are as follows: licensed nurse practitioners must complete a high school degree and an 1800-hour training program, registered nurses must complete an additional year of education beyond high school (CEGEP) and a 2100-hour job training program, and nurse clinicians must complete a three year bachelor degree in Nursing Science. The wage profile also differs widely across the three occupations. The pay differentials are small for new workers but increase substantially with experience, so that a nurse clinician with at least 14 years in the profession has a wage that 80 percent higher than that of a similarly experienced licensed nurse practitioner (FIQ, 2020).^8^ Despite the differences in educational requirements and wages, all three occupations have overlapping job descriptions and responsibilities. Workers in each occupation are directly involved in patient care and common daily tasks include monitoring patient vital signs, administering treatments, providing comfort and hygiene to patients, and interacting with patient family members and friends (Avenir Santé Québec, 2020).

The dataset provides detailed health information on employees. For each worker, we observe whether she had a contact with a COVID-19 positive person in the workplace (either patient or coworker). For workers with a COVID-19 contact, there is information about whether exposure lasted more than 10 minutes and whether the exposure was deemed to be high-risk.^9^

We also have information on diagnostic testing of employees. For each worker, we observe whether a diagnostic test was conducted and the results of the test. We use this information to construct various measures of testing: indicators for whether a worker was ever tested and whether the worker ever tested positive. The dataset also provides information on self-assessed COVID-19 symptoms, which were reported by all nurses regardless of testing. We use this information to construct an indicator for whether a worker ever had COVID-19 symptoms.

The dataset contains information on worker demographics and workplace characteristics. We observe the age, gender, and 3-digit ZIP code of home residence for employees. The data identify the healthcare institution of employment – hospital, nursing home facility, or local community healthcare facility – and the three digit ZIP code of work location.

## 3 Empirical strategy

Our estimation strategy compares health outcomes across nurses of different SES that did or did not have contact with a COVID-19 positive case in the workplace. We estimate the following equation:

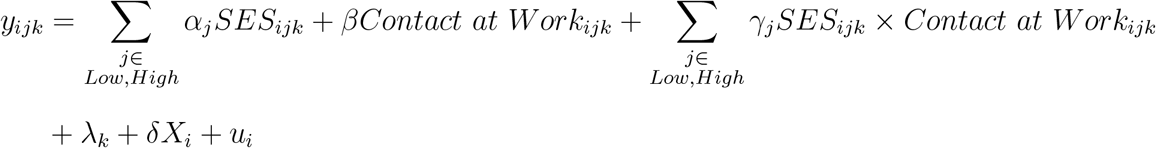

where *y*_*ijk*_ denotes the outcome for nurse *i*, who works in one of three occupations *j* (Licensed Nurse Practitioner, Registered Nurse, or Nurse Clinician) in institution *k* (hospital, CHSLD, or CLSC). The variables *SES*_*ji*_ are indicators for either Low-SES or High-SES. *Contact at Work*_*i*_ is an indicator for a workplace COVID-19 contact. The term *λ*_*k*_ denotes a vector of healthcare institution *×* 3-digit ZIP code interaction effects to allow for within-institution differences in workplace risks that might vary across worksites throughout the city. The term *X*_*i*_ represents a set of covariates for individual age and 3-digit ZIP code of residence to allow for outcomes to vary across individuals of different age groups and different non-workplace environmental conditions. Standard errors are clustered at the 3-digit ZIP code of the workplace to allow for within-group correlation in outcomes.^10^

The coefficients *α*_*Low*_ and *α*_*High*_ identify differences in outcomes across SES groups for individuals that did not have a workplace COVID-19 contact. These effects capture the underlying differences in health outcomes across SES groups that were unrelated to workplace risk. The coefficient *β* identifies the impact of a workplace COVID-19 contact on health outcomes among Mid-SES workers.

The main coefficients of interest, *γ*_*Low*_ and *γ*_*High*_, identify the extent to which workplace COVID-19 exposure differentially affected outcomes for different SES workers. Because these estimates are based on relative differences across exposed versus non-exposed nurses of different SES status, the analysis holds constant all non work sources of COVID-19 risk, thereby isolating the contribution of the workplace to the health gradient. These estimates are derived from comparisons across workers in the same institutions at the same work location, thereby capturing the extent to which the effects of a common workplace risk were borne differently across SES groups.

## 4 Results

To motivate the empirical analysis, we report summary statistics across the the three categories of nurses: Low-SES, Mid-SES and High-SES. Table 1 (Panel A) shows a large and statistically significant SES gradient in workplace exposure, with the highest rates among Low-SES nurses. Conditional on workplace contact, however, the three groups appear to have faced similar levels of risk, as measured by both duration of exposure and whether the contact was deemed high-risk.

**Table 1:**
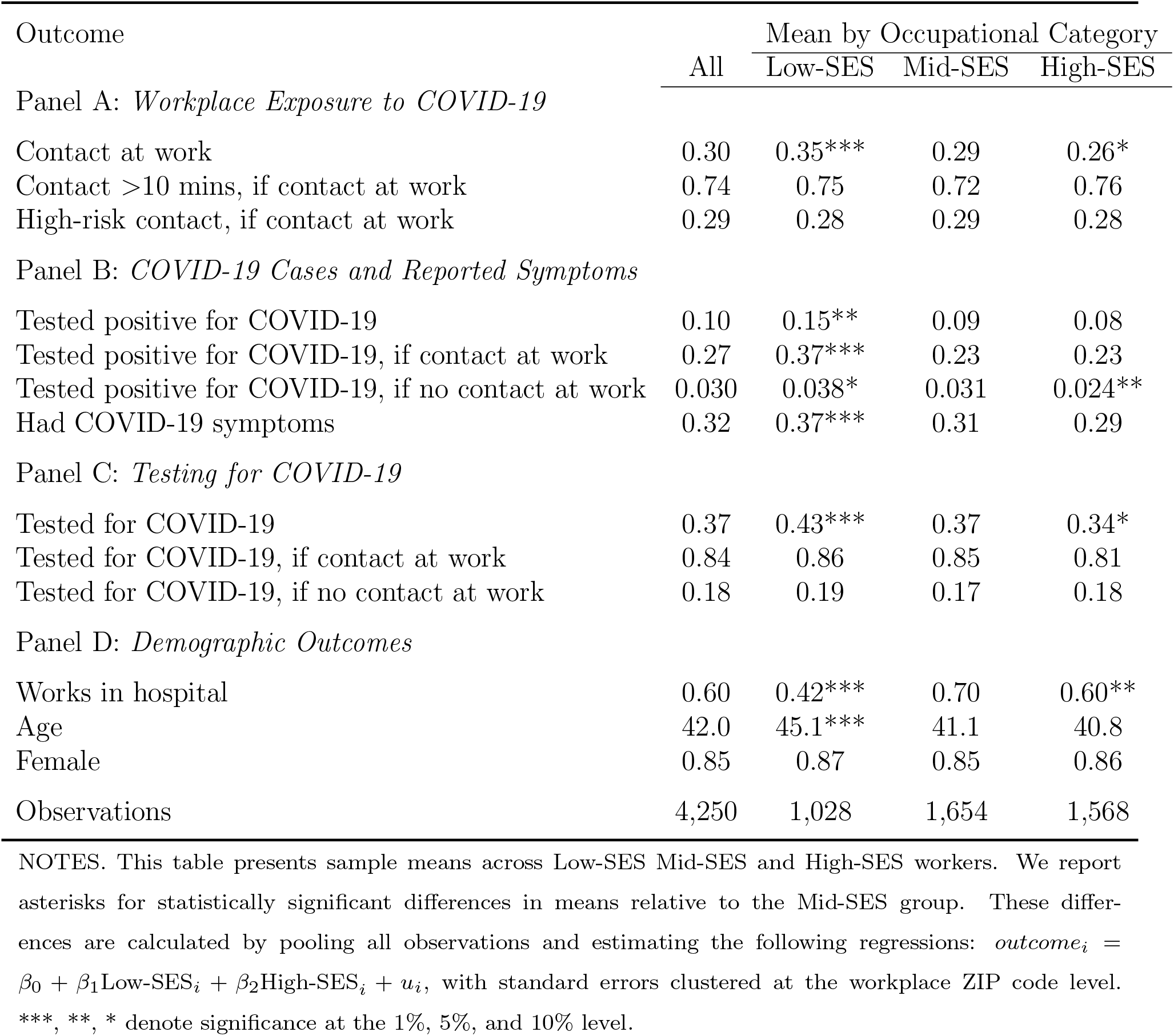
Summary Statistics.

| Outcome | All | Mean by Occupational Category |  |  |
| --- | --- | --- | --- | --- |
|  |  | Low-SES | Mid-SES | High-SES |
| Panel A: *Workplace Exposure to COVID-19* |  |  |  |  |
| Contact at work | 0.30 | 0.35*** | 0.29 | 0.26* |
| Contact >10 mins, if contact at work | 0.74 | 0.75 | 0.72 | 0.76 |
| High-risk contact, if contact at work | 0.29 | 0.28 | 0.29 | 0.28 |
| Panel B: *COVID-19 Cases and Reported Symptoms* |  |  |  |  |
| Tested positive for COVID-19 | 0.10 | 0.15** | 0.09 | 0.08 |
| Tested positive for COVID-19, if contact at work | 0.27 | 0.37*** | 0.23 | 0.23 |
| Tested positive for COVID-19, if no contact at work | 0.030 | 0.038* | 0.031 | 0.024** |
| Had COVID-19 symptoms | 0.32 | 0.37*** | 0.31 | 0.29 |
| Panel C: *Testing for COVID-19* |  |  |  |  |
| Tested for COVID-19 | 0.37 | 0.43*** | 0.37 | 0.34* |
| Tested for COVID-19, if contact at work | 0.84 | 0.86 | 0.85 | 0.81 |
| Tested for COVID-19, if no contact at work | 0.18 | 0.19 | 0.17 | 0.18 |
| Panel D: *Demographic Outcomes* |  |  |  |  |
| Works in hospital | 0.60 | 0.42*** | 0.70 | 0.60** |
| Age | 42.0 | 45.1*** | 41.1 | 40.8 |
| Female | 0.85 | 0.87 | 0.85 | 0.86 |
| Observations | 4,250 | 1,028 | 1,654 | 1,568 |

NOTES. This table presents sample means across Low-SES Mid-SES and High-SES workers. We report asterisks for statistically significant differences in means relative to the Mid-SES group. These differences are calculated by pooling all observations and estimating the following regressions: $outcome_i = \beta_0 + \beta_1 \text{Low-SES}_i + \beta_2 \text{High-SES}_i + u_i$ , with standard errors clustered at the workplace ZIP code level. \*\*\*, \*\*, \* denote significance at the 1%, 5%, and 10% level.

Panel B shows that 10 percent of nurses tested positive for COVID-19, substantially higher than the 1 percent city-wide diagnosis rate (SantéMontréal, 2020).^11^ The high rates of COVID-19 infection are driven by work-related risks. Indeed, the COVID-19 diagnosis rate jumps to 27 percent for nurses with a workplace contact.

We also find systematically higher rates of COVID-19 diagnosis among Low-SES workers, particularly among nurses with a workplace contact. Low-SES nurses were 6 percentage points more likely to be diagnosed for COVID-19 and were significantly more likely to report COVID-19 symptoms. The gap in the positive case rate doubles when we restrict attention to nurses with a workplace contact. For nurses without a workplace contact, COVID-19 diagnosis rates were much lower and similar across SES groups.

Panel C shows that 37 percent of nurses were tested for COVID-19 during the sample period.^12^ COVID-19 tests appear to have been directed to higher risk workers: 84 percent of nurses with a workplace contact received a test. There is a monotonically decreasing relationship between testing rates and occupational ranking, although these differences appear to reflect underlying rates workplace COVID-19 contact. Conditional on workplace contact, there are no significant differences in testing rates across SES groups.

Panel D shows that the three groups also differed across several socioeconomic outcomes. Low-SES nurses were somewhat older and more likely to work in non-hospital settings. Importantly, our estimation strategy controls for both individual characteristics and workplace conditions.

Table 2 reports the results of the estimation of equation (1), where the dependent variable is an indicator equal to 1 if and only if the person tested positive. Columns (1) - (4) report estimates of overall differences in diagnosis rates across SES groups. Across the four specifications, we find that consistently higher rates of COVID-19 diagnosis among Low-SES nurses. Because these models control for workplace ZIP code fixed effects, these differences cannot be attributable to cross institution differences in workplace risk. The findings must reflect either within workplace differences in COVID-19 infection risk or various non-work risks or behavioral differences that are not capture by the demographic covariates.

**Table 2:**
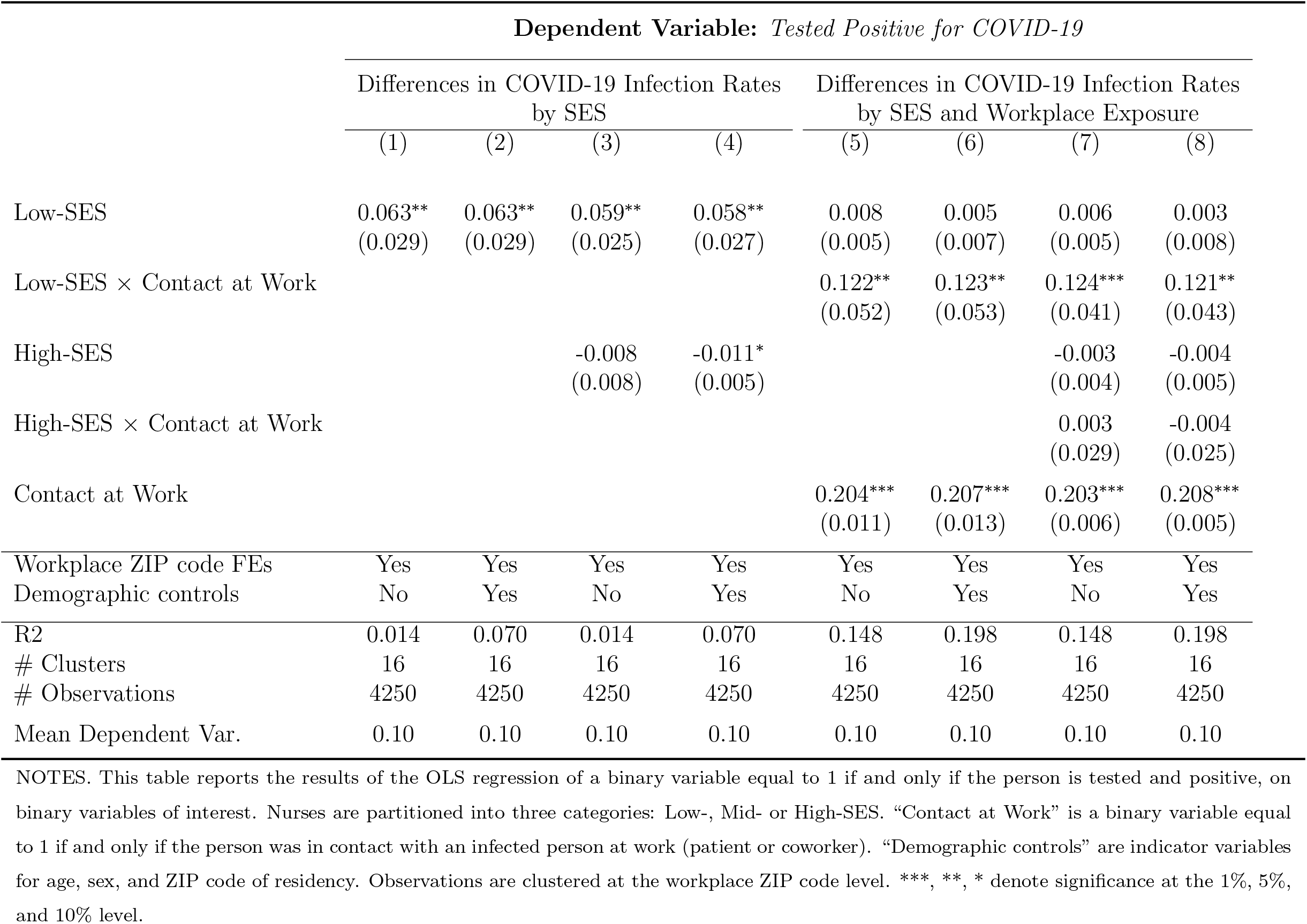
SES Differences in COVID-19 Infections and Workplace Exposure.

|  | Dependent Variable: *Tested Positive for COVID-19* |  |  |  |  |  |  |  |
| --- | --- | --- | --- | --- | --- | --- | --- | --- |
|  | Differences in COVID-19 Infection Rates<br>by SES |  |  |  | Differences in COVID-19 Infection Rates<br>by SES and Workplace Exposure |  |  |  |
|  | (1) | (2) | (3) | (4) | (5) | (6) | (7) | (8) |
| Low-SES | 0.063**<br>(0.029) | 0.063**<br>(0.029) | 0.059**<br>(0.025) | 0.058**<br>(0.027) | 0.008<br>(0.005) | 0.005<br>(0.007) | 0.006<br>(0.005) | 0.003<br>(0.008) |
| Low-SES × Contact at Work | | | | | 0.122**<br>(0.052) | 0.123**<br>(0.053) | 0.124***<br>(0.041) | 0.121**<br>(0.043) |
| High-SES |  |  | -0.008<br>(0.008) | -0.011*<br>(0.005) |  |  | -0.003<br>(0.004) | -0.004<br>(0.005) |
| High-SES × Contact at Work | | | | | | | 0.003<br>(0.029) | -0.004<br>(0.025) |
| Contact at Work |  |  |  |  | 0.204***<br>(0.011) | 0.207***<br>(0.013) | 0.203***<br>(0.006) | 0.208***<br>(0.005) |
| Workplace ZIP code FEs | Yes | Yes | Yes | Yes | Yes | Yes | Yes | Yes |
| Demographic controls | No | Yes | No | Yes | No | Yes | No | Yes |
| R<sup>2</sup> | 0.014 | 0.070 | 0.014 | 0.070 | 0.148 | 0.198 | 0.148 | 0.198 |
| # Clusters | 16 | 16 | 16 | 16 | 16 | 16 | 16 | 16 |
| # Observations | 4250 | 4250 | 4250 | 4250 | 4250 | 4250 | 4250 | 4250 |
| Mean Dependent Var. | 0.10 | 0.10 | 0.10 | 0.10 | 0.10 | 0.10 | 0.10 | 0.10 |

NOTES. This table reports the results of the OLS regression of a binary variable equal to 1 if and only if the person is tested and positive, on binary variables of interest. Nurses are partitioned into three categories: Low-, Mid- or High-SES. "Contact at Work" is a binary variable equal to 1 if and only if the person was in contact with an infected person at work (patient or coworker). "Demographic controls" are indicator variables for age, sex, and ZIP code of residency. Observations are clustered at the workplace ZIP code level. \*\*\*, \*\*, \* denote significance at the 1%, 5%, and 10% level.

Columns 5-8 report the results from our main specification that interact occupational status with workplace contact. The results demonstrate that the underlying differences in positive test rates across groups were driven entirely by workplace contact. The interaction effects for Low-SES are positive and statistically significant across the various specification, implying that workplace contact differentially increased the probability of testing positive among Low-SES nurses. In contrast, the main effects for Low-SES workers are all small and statistically insignificant, indicating no systematic differences in infection rates among individuals who were not exposed in the workplace. Similarly, we find no significant effects for High-SES nurses regardless of workplace exposure.^13^

The interaction effects reported in columns 5-8 are large in magnitude. Our preferred estimates (col. 6) imply that workplace-related infection rates were 53% (= 0.123*/*0.23) higher for Low-SES nurses relative to the other two groups. The findings are consistent with the underlying differences in workplace-related COVID-19 diagnoses reported in Table 1. Notably, the regression results establish that these differentials persist, even after we account for cross-institution differences in infection risk and individual demographic factors.

One potential explanation for the observed patterns in Table 2 is differential testing rates across SES workers. For example, if Low-SES nurses were more likely to be tested following a workplace contact, the previous estimates could partly reflect lower rates of undiagnosed infection among this group of workers. In practice, decisions over employee testing were made at the institutional level, following the guidelines established by the provincial health authorities. These guidelines made no mention of nurses’ occupational standing, so it is unlikely that testing rates should differ across groups, *conditional* on underlying workplace risk. Nevertheless, we assess the extent to which group-specific testing policies may be driving the baseline findings. Table 3 (cols. 1-2) shows no significant difference in the testing rates for Low-SES workers, regardless of workplace contact. The main interaction effects are positive, but small in magnitude, and statistically insignificant. The main results for COVID-19 diagnosis are similar to the baseline findings when we restrict the sample to individuals who received a COVID-19 tests (cols. 3-4). Similarly, we estimate a significant workplace-related SES gradient in the probably of reporting COVID-19 symptoms, *regardless* of whether a diagnostic test was conducted (cols. 5-6). Taken together, these results strongly suggest that the observed differentials in positive COVID-19 cases across nurses reflect actual differences in the likelihood of workplace-related infections and were not driven by SES-based differences in testing.

**Table 3:** SES Differences in COVID-19 Testing.

| Dependent Variable: | Tested<br>for COVID-19 |  | COVID-19<br>positive,<br>if tested =1 |  | Reported<br>COVID-19<br>symptoms |  |
| --- | --- | --- | --- | --- | --- | --- |
|  | (1) | (2) | (3) | (4) | (5) | (6) |
| Low-SES | 0.009<br>(0.019) | 0.006<br>(0.026) | 0.042<br>(0.034) | 0.045<br>(0.041) | -0.001<br>(0.018) | -0.006<br>(0.025) |
| Low-SES × Contact at Work | 0.014<br>(0.039) | 0.019<br>(0.038) | 0.100**<br>(0.034) | 0.097**<br>(0.033) | 0.061*<br>(0.029) | 0.068**<br>(0.031) |
| Contact at Work | 0.657***<br>(0.017) | 0.653***<br>(0.015) | 0.122***<br>(0.016) | 0.113***<br>(0.015) | 0.542***<br>(0.017) | 0.543***<br>(0.020) |
| Workplace ZIP code FEs | Yes | Yes | Yes | Yes | Yes | Yes |
| Demographic controls | No | Yes | No | Yes | No | Yes |
| Restrict sample to<br>tested workers |  |  | Yes | Yes |  |  |
| R2 | 0.393 | 0.429 | 0.051 | 0.193 | 0.308 | 0.351 |
| # Clusters | 16 | 16 | 16 | 16 | 16 | 16 |
| # Observations | 4250 | 4250 | 1587 | 1587 | 4250 | 4250 |
| Mean Dependent Var. | 0.37 | 0.37 | 0.27 | 0.27 | 0.32 | 0.32 |

NOTES. This table reports the results of the OLS regression of a binary variable equal to 1 if and only if the person is tested and positive, on binary variables of interest. Nurses are partitioned into three categories: Low-, Mid- or High-SES. “Contact at Work” is a binary variable equal to 1 if and only if the person was in contact with an infected person at work (patient or coworker). “Demographic controls” are indicator variables for age, sex, and ZIP code of residency. Observations are clustered at the workplace ZIP code level. \*\*\*, \*\*, \* denote significance at the 1%, 5%, and 10% level.

The absence of SES-based testing differentials is unsurprising, since employee occupational standing did not directly factor into the provincial testing guidelines. Nevertheless, these patterns are striking, given that a primary goal of workplace testing was to identify the maximum number of COVID-19 cases among workforce. According to this objective, tests should be allocated disproportionately to all identifiable group with higher underlying risk of COVID-19 infection. Instead, we find that Low-SES workers had significantly higher rates of workplace-related infection but were no more likely to be tested, suggesting that a reallocation of tests to Low-SES workers may have reduced the rates of undiagnosed infection.

What explains the higher rates of COVID-19 infection among Low-SES workers? One possibility is that these nurses were given tasks that placed them at higher risk of infection. For example, they may have been more likely to spend longer periods of time caring for COVID-19 positive patients or may have been assigned to patients that posed higher risks to caregivers. In Table 4, we assess whether different SES workers faced different levels of risk in the workplace. In columns 1-3 we explore whether there were systematic differences in the duration of workplace COVID-19 contact. Overall, Low-SES workers were more likely to experience an extended COVID-19 contact (col. 1). Conditional on contact, however, we find no significant differences in the length of time spent with COVID-positive patients (col. 2). Moreover, the inclusion of a controls for contact duration has almost no impact on the main interaction estimates (col. 3). Similarly, we find no evidence that Low-SES had higher rates of high-risk workplace contact, and controlling for high-risk contact does not alter the main estimates (cols. 4-6).^14^

**Table 4:**
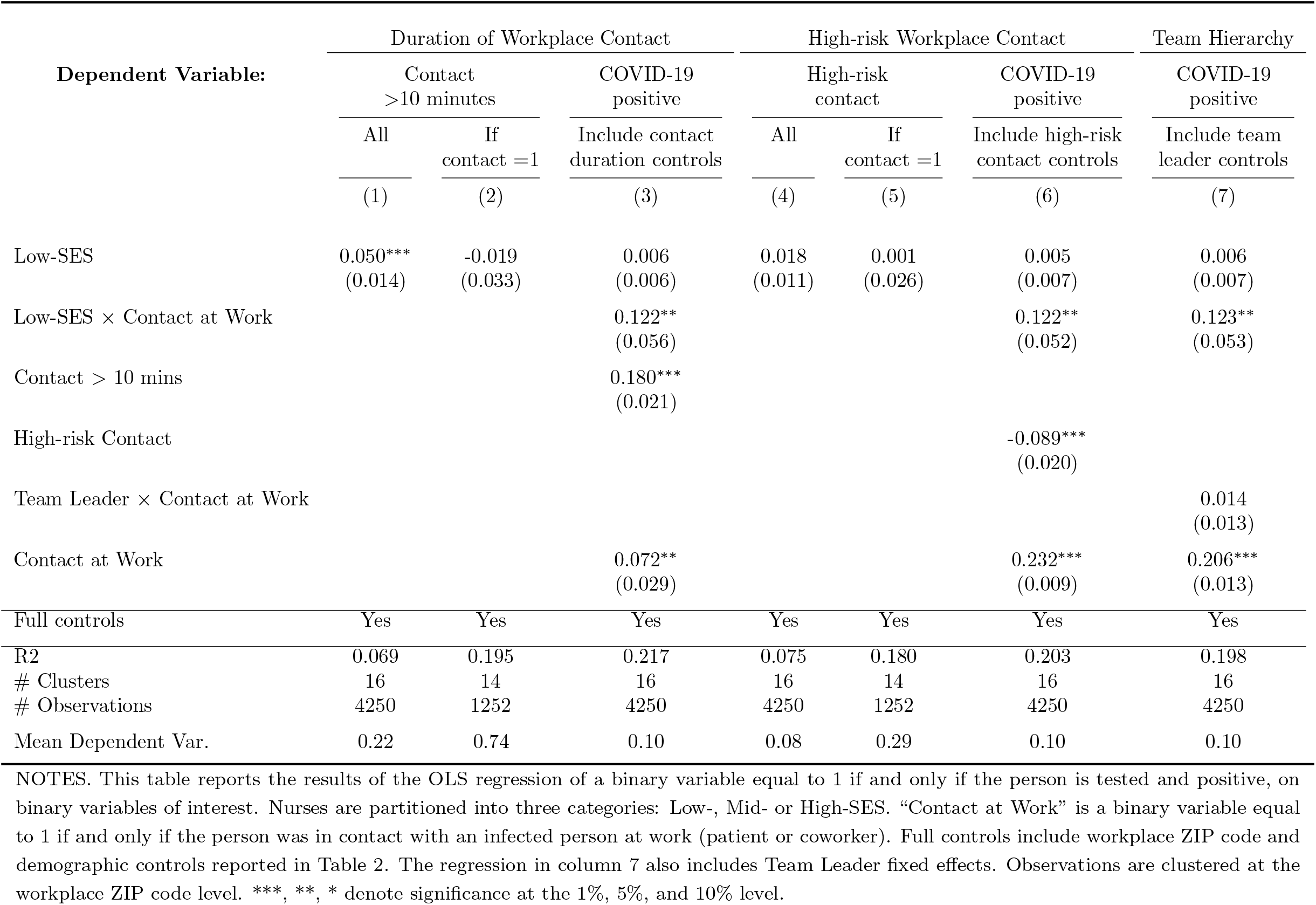
SES Differences in Workplace COVID-19 Infection Risk.

In the final column of Table 4, we assess whether the SES gradient can be attributed to the workplace hierarchy. We estimate a generalized version of equation (1), allowing the effect of workplace exposure to vary according to whether a nurse was the team leader. This framework allows us to test whether team leaders, who were more likely to Mid- or High-SES nurses, assigned themselves less risky tasks. We estimate a positive (albeit statistically insignificant) interaction effect for team leaders, suggesting that head nurses were somewhat more likely to assign *themselves* to higher-risk patients. The inclusion of these covariates has little effect on the main interaction terms.

Taken together, the results in Tables 4 show no evidence that Low-SES nurses spent more time with COVID-19 patients, or were assigned to patients at higher risk of transmitting the virus. Instead, the results must reflect more subtle differences in the daily tasks performed by each group that placed some groups at systematically higher risk of infection.

To conclude the analysis, we explore heterogeneity in effects across different healthcare institutions. In Quebec, like many other jurisdictions, there were substantial differences in the effectiveness of the COVID-19 response across different healthcare institutions.^15^ The outbreak was particularly acute in elder living institutions (CHSLDs), which accounted for more than 60 percent of province-wide deaths during the first wave. A widely publicized report by the Quebec ombudsman noted that “CHSLDs were a blind spot in preparing for the pandemic, with efforts massively concentrated on hospitals (Rinfret, December 10, 2020).” The report documented that there was a “lack of infection prevention and control culture in CHSLDs” and that “personal protective equipment was insufficient and unequally distributed.” Motivated by this evidence, we assess the extent which the SES workplace infection gradient differed across hospital and non-hospital settings.

Table 5 reports the results from regressions that allow the Low-SES interaction effect to differ across hospital and non-hospital healthcare institutions. The estimates for COVID-19 diagnosis are positive and statistically in both hospital and non-hospital settings (cols. 1-2), suggesting that the SES gradient existed in both workplace settings. The estimates are more than twice as large in non-hospital institutions. These findings do not appear to have been driven by differential testing policies (cols. 5-6) and the differential effects remain even when we restrict the sample to tested workers (cols. 3-4).^16^ Taken together, these findings suggest that the costs of PPE shortages and poor preparedness in many non-hospital healthcare facilities were disproportionately borne by Low-SES workers.

**Table 5:** SES Differences in Workplace COVID-19 Infection Risk Across Healthcare Institutions.

| Dependent Variable: | COVID-19 positive |  | COVID-19 positive,<br>if tested = 1 |  | Tested for<br>COVID-19 |  |
| --- | --- | --- | --- | --- | --- | --- |
|  | (1) | (2) | (3) | (4) | (5) | (6) |
| Low-SES × Contact at Work × Hospital | 0.062***<br>(0.011) | 0.068***<br>(0.013) | 0.030*<br>(0.016) | 0.044*<br>(0.023) | 0.012<br>(0.017) | 0.011<br>(0.016) |
| Low-SES × Contact at Work × Non-hospital | 0.155***<br>(0.051) | 0.151**<br>(0.054) | 0.137***<br>(0.043) | 0.120**<br>(0.050) | 0.022<br>(0.051) | 0.031<br>(0.048) |
| Workplace ZIP code FEs | Yes | Yes | Yes | Yes | Yes | Yes |
| Demographic controls | No | Yes | No | Yes | No | Yes |
| Restrict sample to tested workers |  |  | Yes | Yes |  |  |
| R<sup>2</sup> | 0.150 | 0.200 | 0.055 | 0.197 | 0.393 | 0.430 |
| # Clusters | 16 | 16 | 16 | 16 | 16 | 16 |
| # Observations | 4250 | 4250 | 1587 | 1587 | 4250 | 4250 |
| Mean Dependent Var. | 0.10 | 0.10 | 0.27 | 0.27 | 0.37 | 0.37 |

NOTES. This table reports the results of the OLS regression of a binary variable equal to 1 if and only if the person is tested and positive, on binary variables of interest. Nurses are partitioned into three categories: Low-, Mid- or High-SES. "Contact at Work" is a binary variable equal to 1 if and only if the person was in contact with an infected person at work (patient or coworker). "Demographic controls" are indicator variables for age, sex, and ZIP code of residency. All models include double-interactions between Low-SES, hospital and non-hospital dummies as well as the direct effect of Contact at Work. Observations are clustered at the workplace ZIP code level. \*\*\*, \*\*, \* denote significance at the 1%, 5%, and 10% level.

## 5 Assessing the Risks of COVID-19 Transmission in the Workplace

Policymakers in many countries have implemented partial lockdowns in which many businesses have been required to shutdown or transition to home-based work to reduce the spread of the COVID-19 pandemic. These decisions have been highly controversial, and there has been considerable debate both their scope of workplace shutdowns and the targeted industries. One reason for the controversy is the large uncertainty regarding the actual risks of workplace infection and how these risks vary across occupations.

In an effort to address these concerns, researchers have developed tools to assess the relative risk of workplace infection as a function of the specific daily tasks involved with various jobs. These models combine detailed data on specific job characteristics and tasks from the O*NET dataset with assessments from health experts on the associated risks of viral transmission. For example, a job that requires daily face-to-face discussions would be classified as higher risk than one that requires only weekly interactions. The vector of these various job attributes can then be combined to construct an index of viral transmission risk at the industry/occupation level. These tools have gained influence and have been applied in a number of studies.

A challenge for the ‘task-based’ approach to measuring workplace transmission risk is that the various indices have not been validated with *actual* COVID-19 infection rates. As a result, we do not know the extent to which the risks of transmission identified by public health experts align with actual rates of transmission. This concern is particularly relevant for the current pandemic, in which knowledge of sources of transmission continues to evolve.

We compare our estimates of relative transmission risk across licensed practitioner nurses and registered nurses to those implied by a standard occupational ‘task-based’ index (Baylis et al., 2020*a*).^17^ We corroborate the ranking of these occupational groups: licensed practical nurses face higher risk of COVID-19 infection according to the ‘task-based’ index. Nevertheless, the magnitude of the differences in risk do not align. Our preferred estimates imply that the former group had a 50 percent higher risk of workplace-related infection. In contrast, the gap in infection risk on the 100 point ‘task-based’ index is 4 points, less than one third of the cross-occupation standard deviation in risk. Combining our estimated differences in infection risk with the occupation/industry risk index, we calculate that an implausible 98 percent of jobs carry zero risk of transmission risk.^18^ We also found that the occupational differences in workplace infection risk were three times higher in non-hospital settings that were not identifiable in occupational-based indices.

These findings demonstrate the need to interpret with caution ‘task-based’ occupational measures of transmission risk. There may be subtle differences across occupations that are not easily quantified but that may importantly impact transmission risk. Moreover, our finding that the SES gradient differs substantially across different institutions highlights the crucial role of specific worksite conditions for disease spread. Our results demonstrate the critical need for data on *actual* COVID-19 infection rates across a broader set of workers to better understand how workplace conditions contribute to disease transmission.

## 6 Conclusion

The positive relationship between socioeconomic status and health has been extensively documented. Many national governments have set ambitious targets to reduce these “health inequalities” (WHO, 2008; U.S. DHHS, 2010). The unequal impacts of ongoing coronavirus pandemic have only increased the urgency of this issue, although the effectiveness of government efforts will hinge on successfully identifying and addressing the underlying causes of the gradient. Our study provides new evidence on the sources of health disparities among frontline healthcare workers during the COVID-19 pandemic. We find that large SES-based differences in COVID-19 infection rates. This gradient was driven entirely by work-related contact, and we find no differences infection rates among individuals who were not exposed to COVID-19 in the workplace.

We uncover wide differences in work-related COVID-19 infections, even among individuals within narrowly defined occupational categories who worked in the same healthcare institution. These differences are not easily captured by standard measures of workplace risk or ‘task-based’ indices of disease transmission. Instead, our results show how subtle differences in workplace conditions and job tasks can interact to generate large SES health disparities.

## Data Availability

Data for the study can be obtained through an application process to the CIUSSS and the CISSS for the Greater Montreal Area.

## A Appendix (For Online Publication)

**Table A.1:** SES Differences in COVID-19 Infections and Workplace Exposure.

|  | Dependent Variable: *Tested Positive for COVID-19* |  |  |  |  |  |  |  |
| --- | --- | --- | --- | --- | --- | --- | --- | --- |
|  | Differences in COVID-19 Infection Rates<br>by SES |  |  |  | Differences in COVID-19 Infection Rates<br>by SES and Workplace Exposure |  |  |  |
|  | (1) | (2) | (3) | (4) | (5) | (6) | (7) | (8) |
| Low-SES Nurse | 0.063**<br>(0.028) | 0.063*<br>(0.030) | 0.059**<br>(0.024) | 0.058*<br>(0.029) | 0.008<br>(0.005) | 0.005<br>(0.008) | 0.006<br>(0.006) | 0.003<br>(0.009) |
| Low-SES × Contact at Work | | | | | 0.122**<br>(0.050) | 0.123**<br>(0.054) | 0.124***<br>(0.039) | 0.121**<br>(0.045) |
| High-SES Nurse |  |  | -0.008<br>(0.008) | -0.011<br>(0.008) |  |  | -0.003<br>(0.004) | -0.004<br>(0.008) |
| High-SES × Contact at Work | | | | | | | 0.003<br>(0.029) | -0.004<br>(0.028) |
| Contact at Work |  |  |  |  | 0.204***<br>(0.011) | 0.207***<br>(0.016) | 0.203***<br>(0.005) | 0.208***<br>(0.011) |
| Workplace zipcode FE | Yes | Yes | Yes | Yes | Yes | Yes | Yes | Yes |
| Full controls | No | Yes | No | Yes | No | Yes | No | Yes |
| R2 | 0.014 | 0.070 | 0.014 | 0.070 | 0.148 | 0.198 | 0.148 | 0.198 |
| # Observations | 4247 | 4247 | 4247 | 4247 | 4247 | 4247 | 4247 | 4247 |
| Mean Dependent Var. | .100 | .100 | .100 | .100 | .100 | .100 | .100 | .100 |

NOTES. This table reports the results of the OLS regression of a binary variable equal to 1 if and only if the person is tested and positive, on binary variables of interest. Nurses are partitioned into three categories: Low-, Mid- or High-SES. "Contact at Work" is a binary variable equal to 1 if and only if the person was in contact with an infected person at work (patient or coworker). "Demographic controls" are indicator variables for age, sex, and ZIP code of residency. Observations are two-way clustered at the workplace and residency ZIP code level, following the approach of Cameron, Gelbach and Miller \(2011\). \*\*\*, \*\*, \* denote significance at the 1%, 5%, and 10% level.

## Footnotes

1 By July 1, the total number of diagnosed COVID-19 cases per 1,000 residents was 16 in Montreal, comparable to the rates in Baltimore (13), Washington D.C. (15), Philadelphia (17), and Boston (20), although lower than those in New York City (27) (SantéMontréal, 2020; Maryland Dept. of Health, 2020; Government of the District of Columbia, 2020; Philadelphia Dept. of Public Health, 2020; Massachusetts Dept. of Public Health, 2020; NY State Dept. of Health, 2020).

2 For example, among nurses with at least 14 years of experience, the hourly wage gap between NCs and LPNs is 80 percent (FIQ, 2020).

3 The decision to test an employee was made at the institutional level following guidelines provided by the Institut National de SantéPublique du Québec (INSPQ). Occupation did not factor directly into this determination, although cross-group testing differentials could arise as a consequence of differences in workplace risks.

4 See Li et al. (2020) for the rates of asymptomatic COVID-19 infection.

5 A series of influential papers based on the Whitehall study of male British Civil Servants argued that job stress and lack of control over job tasks among lower status workers contributed to poor health (Marmot, Kogevinas and Elston, 1987; Marmot et al., 1991).

6 The dataset also information on orderlies. There is some information on doctors in these institutions, although most are classified as self-employed and not administered by the same Human Resources department.

7 Roughly 95 percent of Quebec nurses are unionized, and almost all are represented by the Fédération Interprofessionnelle de la Santéde Québec (FIQ).

8 During the pandemic, the Quebec government provided wage increases to frontline healthcare workers that varied across institutions and depending whether there was a confirmed case at the worksite (Harris, C., May 7, 2020). More generally, wages also varied based on overtime work and additional certifications.

9 The ‘high-risk’ classification was made at the discretion of hospital administrators based on a variety of factors including proximity to the COVID-19 positive individual, the use of PPE, and duration of contact.

10 We also estimate two-way clustered standard errors that allow for correlation within both workplace and home ZIP code addresses, following the approach of Cameron, Gelbach and Miller (2011).

11 These disparities may reflect a combination of both the higher risk of infection among healthcare workers (Wu and McGoogan, 2020), and the high rates of testing that lowered the probability of undiagnosed infection (Benatia, Godefroy and Lewis, 2020*a,b*).

12 In comparison, just 7 percent of the general population in the population had been tested by the end of the sample period (INSPQ, 2020).

13 In Table A.1 we report results from models with two-way clustered standard errors to allow for correlation in outcomes within both workplace and home ZIP code address, following the approach of Cameron, Gelbach and Miller (2011). Statistical inference is unaffected.

14 The negative direct effect of ‘High-risk Contact’ is likely a result of differential testing rates among workers whose exposure was deemed ‘high-risk’ (results available upon request). Differences in COVID-19 diagnosis rates across workers with high- and low-risk contacts reflect *both* average COVID-19 infection rates and differences in testing rates across each group. To the extent that the positivity rate decreases with the fraction of the population tested, this second selection effect may dominate.

15 For details on PPE shortages in U.S. elder care facilities see Murray (2020); Holroyd-Leduc and Laupacis (2020).

16 In these regressions, we are unable to identify the main effect of workplace exposure on COVID-19 diagnosis, since *overall* testing rates differed across hospital and non-hospital institutions.

17 Nurse clinicians were not separately identified in the O*NET data.

18 This large share is due to the implied steep decline in COVID-19 infection rates for jobs that had lower rankings on the index.

